# The Adequacy of ANC services received and associated factors among Women of Reproductive Age in Tanzania

**DOI:** 10.1101/2024.03.20.24304603

**Authors:** Jovin R. Tibenderana, Sanun Ally Kessy, Dosanto Felix Mlaponi, John Elyas Mtenga, Jomo Gimonge, Ndinagwe Lloyd Mwaitete, Fabiola V Moshi

## Abstract

**Background:** ANC continues to be a vital and crucial component of care for expectant mothers and their unborn children, not only by lowering maternal mortality but also perinatal deaths and connecting them to postnatal treatment. There are limited information about the adequate Antenatal Care (ANC) services coverage, therefore this study aimed at examining the proportion of ANC services coverage, distribution pattern and its associated factors obtained by women of reproductive age (WRA) in Tanzania.

**Methods:** This was analytical cross-sectional study among women of reproductive age in Tanzania, utilizing data from the Demographic and Health Surveys (DHS) 2022. Analysis considered the complex survey design through application of weights, clustering and strata. Modified Poisson regression models estimated the factors associated with adequate reception of ANC services among WRA in Tanzania. All analyses were performed in STATA software version 18.

**Results:** The proportion of WRA who had received adequate ANC component coverage was 41%. Distribution of ANC contents ranged from 0.5% to 41% for none to seven services respectively. Almost all women (96.1%) had their baby’s heartbeat checked. Various factors such as education, wealth index, age, residence, number of ANC visits and birth order were significantly associated with uptake of ANC services among WRA in Tanzania.

**Conclusion:** The overall findings suggest a notable disparity in the utilization of ANC services, as fewer than half of women of reproductive age (WRA) are receiving sufficient ANC coverage. Therefore, integrated approaches involving health care facilities and communities, innovative strategies targeting young adults, and strengthening the policy of four or more ANC visits with regular monitoring and data collection are recommended to improve ANC access, utilization, and alignment with WHO guidelines.

## Background

ANC has remained a critical and pivotal point in pregnant mothers and their babies not only by reducing maternal mortality but also perinatal deaths together with linkage to postnatal care (1,2). Globally 287,000 maternal deaths occur, out of which 70% occurred in sub-Saharan Africa with the highest Maternal Mortality Ratio (MMR) of 545 per 100, 000 livebirths in 2020 (3). In Tanzania, the maternal mortality ratio is estimated at 104 deaths per 100,000 live births (4). The slow reduction of maternal mortality in Tanzania continues to threaten the achievement of the global agendas of reducing maternal mortality to 70 deaths per 100,000 live births by 2030 as per Sustainable Development Goal 3.1(5)(6).

Inadequate uptake of the ANC services and variation around low and middle income countries has led to stagnation in reduction of maternal mortality rate in various areas of the country and pregnancy related complications such as preeclampsia and GDM also resulting into poor childbirth outcomes(1,7,8).

In low- and middle-income countries, increased ANC coverage and high-quality healthcare can prevent 71% of neonatal deaths, 33% of stillbirths, and 54% of maternal deaths(9). Women who received more visits and started ANC in first trimester are more likely to receive all routine services (10). Young women and those with history of unintended pregnancy were less likely to receive sufficient component as well as adequate ANC (11). Mothers who had less than four visits and who had unintended pregnancy were less likely to receive adequate ANC component(12). Benage et al., highlighted that Regional difference plays a role in adequate antenatal care component(13).

ANC includes the following services: risk identification; prevention and management of pregnancy-related or concurrent diseases; and health education and promotion. The proportion of women with overall adequate ANC ranged from 9.6% in Uganda (10), 11% and 19.6% in Ethiopia (11,12) to 21% in Bangladesh (14). Studies from Tanzania, Kenya, and Rwanda have revealed inadequate counseling during ANC, with just 68% of women receiving advice on birth preparedness, 55% on risk indications, 41% on family planning after childbirth, and 17% on nutrition (6). In Tanzania inadequate ANC was reported in a rural district in eastern-central, where hemoglobin was measured in 22 % to 37 % of women and Blood pressure measurement was most common service provided to about 69% and 87% of women attending ANC (14–17) with the lowest coverage being urine sample taken (18,19).

Factors associated with adequate/ inadequate reception of ANC services among WRA include rural/urban residence (9,15,20,21) education level(9,15,21–23), household wealth (15,21), use of private ANCs (18,21,23), high risk pregnant women (21), number of ANC check-ups made by a woman during her pregnancy (15), timely ANC initiation(24) age at marriage (15), maternal age, partner’s encouragement to attend ANC clinic (9) and having no current health complications (23).

There is a limited number of researches on adequate ANC services coverage and therefore this study aims at examining the proportion of ANC services coverage, distribution pattern and its associated factors obtained by WRA in Tanzania. The results of this study will provide insights to policy makers about the different public health strategies to increase the coverage of adequate ANC services among pregnant women

## Methods and materials

### Study design and period

This was analytical cross sectional study conducted by utilizing data collected from Tanzania Demographic and Health Survey (TDHS) in 2022(4). Tanzania is the largest country in East Africa, covering 940,000 square kilometers, 60,000 of which are inland and water and its population in 2022 was estimated to be 61,741,120 with an annual population growth rate of 3.2% (4)(25) .

### Data source/extraction

Once authorization was obtained via an online request outlining the purpose of the research, the Measure Demographic and Health Surveys (DHS) website was accessed to obtain the data. (https://dhsprogram.com/)

### Population of the study

The study included all women of reproductive ages 15-49 years who have given birth in the past 2 years prior on survey who had at least one ANC visit for their last child all over Tanzania.

### Eligibility criteria

All women of reproductive age who gave birth during the five years prior to the survey and who were discovered in the designated clusters at least one night before the data collection period were included in the study; women without an ANC visit or with an unknown ANC visit date were excluded. In consideration of this, 5,263 weighted samples of women in reproductive age were included.

### Sampling technique

A detailed description of the DHS sampling method can be found in NBS and TDHS (4)(25). To obtain a complete representation of the target population, TDHS uses a two-stage probability sampling technique from a sampling frame that is already in place, the census frame. The sampling technique intends to provide national estimates for urban and rural areas of Tanzania mainland and for Zanzibar.First sampling stage involved selection of primary sampling units (PSUs) with probability proportion to size (PPS) within each stratum. The PSU are Census Enumeration areas (EAs), the PSU also from the survey clusters. In the second stage, households were selected proportionally from each EA by using a systematic sampling method.

### Variables of the study

#### Dependent variable

Binary outcome whether a women receive Adequacy (coded as 1=Yes), or inadequacy (coded as 0=No) of ANC components, this was adopted from previous studies and defined as Adequate if a woman received all 7 selected services during her ANC visit and Inadequate if a woman received 6 or less selected ANC services (26,27).

Women of reproductive age were asked by health care provider as part of your antenatal care during this pregnancy, did a healthcare provider do any of the following: Measure your blood pressure, Take a urine sample, Take a blood sample, Listen to the baby’s heartbeat, talk with you about which foods you should eat, Talk with you about breastfeeding, ask you if you had vaginal bleeding, and their answer was either Yes or No which was coded (1=Yes and 0=No) (4,28).

#### Independent variables

The independent variables were classified as individual and community-level variables. The individual-level variables include maternal age, marital status, educational status of women, household wealth status, Birth order, Employment status, number of ANC Visits, Partner’s education. Whereas the place of residence of the study participants were considered as community-level variables. The detailed information’s about the explanatory factors were presented in table 1

**Table 1:** description of the independent variable’s categorization and definitions.

| Variable | Definitions/categorizations |
| --- | --- |
| Education | Mothers' education was categories into three subcategories in which secondary and primary was combined into one category and coded as follow, No education, Primary/Secondary, or higher. |
| Marital status | Marital status was coded into two categories Married and unmarried (never in union, living with partner, widowed, divorced, no longer living together/separated was group into one category as unmarried). |
| Employment status | Employment status was categories into Not working and working, based on the previous studies working was derived by combining the following variables (legislators, administrators and manager, professionals, technicians and associate professionals, clerks, service workers and shop sales workers, skilled agricultural and fishery worker, craft and related workers, plant and machine operators and assembly, elementary |
|  | occupations, Tanzania peoples defense force, national service, and others) |
| ANC Visits | The total number of ANC visit was ranging from 0 to 16, those who were coded 0 excluded from our since they do not meet inclusion criteria, then visit 1 to 3 was group into one category and coded as < 4 and ANC visit 4 to 16 was group into one category and coded as >4. |
| Wealth index | Wealth index was derived and categorized into three subcategories Poor (poorest and poorer), Middle, Rich (richer and richest) |
| Partner's education | Partner's education was categories into three subcategories in which secondary and primary was combined into one category and coded as follow, No education, Primary/Secondary, or higher. |
| Birth order | Birth order was a count number ranging from 0 to 5, those who were coded 0 excluded from our since they do not meet inclusion criteria however, we regrouped into 1 and 2+. |
| Age categories | To increase the power of the study and based on the previous literature mothers aged was coded into three groups as follows 15_24, 25_34 and 35_49. |
| Residence | Residence was coded as Urban and Rural. |

**Table 1:** ANC services received by socio-demographic characteristics (N=5,263)

|  | **Blood pressure measured** | **Urine sample taken** | **Blood sample taken** | **Baby's heartbeats checked** | **Counselled about maternal diet** | **Counselled about breastfeeding** | **Asked about Vaginal bleeding** |
| --- | --- | --- | --- | --- | --- | --- | --- |
|  | **Yes, n (%)** | **Yes, n (%)** | **Yes, n (%)** | **Yes, n (%)** | **Yes, n (%)** | **Yes, n (%)** | **Yes, n (%)** |
| **Education** |  |  |  |  |  |  |  |
| No education | 663 (16.2) | 590 (15.8) | 906 (18.6) | 963 (18.8) | 586 (15.7) | 586 (15.7) | 581 (16.3) |
| Primary | 2,255 (55.0) | 2,025 (54.1) | 2,698 (55.4) | 2,863 (56.0) | 2,065 (55.4) | 2,065 (55.4) | 1,988 (55.7) |
| Secondary or higher | 1,180 (28.8) | 1,128 (30.1) | 1,267 (26.0) | 1,285 (25.1) | 1,078 (28.9) | 1,078 (28.9) | 1,001 (28.0) |
| **Marital status** |  |  |  |  |  |  |  |
| Unmarried | 1,771 (43.2) | 1,622 (43.3) | 2,059 (42.3) | 2,229 (43.6) | 1,625 (43.6) | 1,625 (43.6) | 1,558 (43.6) |
| Married | 2,327 (56.8) | 2,121 (56.7) | 2,812 (57.7) | 2,882 (56.4) | 2,105 (56.4) | 2,105 (56.4) | 2,012 (56.4) |
| **Employment status** |  |  |  |  |  |  |  |
| Not working | 1,348 (32.9) | 1,258 (33.6) | 1,568 (32.2) | 1,655 (32.4) | 1,219 (32.7) | 1,219 (32.7) | 1,162 (32.6) |
| Working | 2,749 (67.1) | 2,484 (66.4) | 3,303 (67.8) | 3,457 (67.6) | 2,511 (67.3) | 2,511 (67.3) | 2,408 (67.4) |
| **ANC Visits** |  |  |  |  |  |  |  |
| <4 | 1,990 (48.6) | 1,747 (46.7) | 2,511 (51.5) | 2,715 (53.1) | 1,816 (48.7) | 1,816 (48.7) | 1,709 (47.9) |
| >4 | 2,108 (51.4) | 1,996 (53.3) | 2,360 (48.5) | 2,396 (46.9) | 1,914 (51.3) | 1,914 (51.3) | 1,862 (52.1) |
| **Wealth index** |  |  |  |  |  |  |  |
| Poor | 1,426 (34.8) | 1,212 (32.4) | 1,894 (38.9) | 2,030 (39.7) | 1,302 (34.9) | 1,302 (34.9) | 1,257 (35.2) |
| Middle | 766 (18.7) | 675 (18.0) | 925 (19.0) | 996 (19.5) | 701 (18.8) | 701 (18.8) | 689 (19.3) |
| Rich | 1,906 (46.5) | 1,856 (49.6) | 2,052 (42.1) | 2,085 (40.8) | 1,727 (46.3) | 1,727 (46.3) | 1,625 (45.5) |
| **Partner's education** |  |  |  |  |  |  |  |
| No education | 362 (8.8) | 310 (8.3) | 493 (10.1) | 532 (10.4) | 313 (8.4) | 313 (8.4) | 330 (9.2) |
| Primary | 2,706 (66.0) | 2,453 (65.5) | 3,279 (67.3) | 3,461 (67.7) | 2,491 (66.8) | 2,491 (66.8) | 2,383 (66.8) |
| Secondary or higher | 1,029 (25.1) | 980 (26.2) | 1,099 (22.6) | 1,119 (21.9) | 926 (24.8) | 926 (24.8) | 858 (24.0) |
| **Birth order** |  |  |  |  |  |  |  |
| 1 | 2,435 (59.4) | 2,285 (61.1) | 2,791 (57.3) | 2,896 (56.7) | 2,202 (59.0) | 2,100 (58.2) | 2,094 (58.7) |
| 2+ | 1,663 (40.6) | 1,457 (38.9) | 2,081 (42.7) | 2,215 (43.3) | 1,528 (41.0) | 1,511 (41.8) | 1,476 (41.3) |
| **Age categories** |  |  |  |  |  |  |  |
| 15_24 | 1,339 (32.7) | 1,216 (32.5) | 1,639 (33.6) | 1,725 (33.8) | 1,233 (33.1) | 1,233 (33.1) | 1,184 (33.2) |
| 25_34 | 1,830 (44.7) | 1,698 (45.4) | 2,151 (44.1) | 2,258 (44.2) | 1,687 (45.2) | 1,687 (45.2) | 1,589 (44.5) |
| 35_49 | 928 (22.7) | 829 (22.2) | 1,082 (22.2) | 1,128 (22.1) | 810 (21.7) | 810 (21.7) | 797 (22.3) |
| **Residence** |  |  |  |  |  |  |  |
| Urban | 1,432 (35.0) | 1,380 (36.9) | 1,497 (30.7) | 1,493 (29.2) | 1,250 (33.5) | 1,250 (33.5) | 1,168 (32.7) |
| Rural | 2,665 (65.0) | 2,363 (63.1) | 3,374 (69.3) | 3,618 (70.8) | 2,480 (66.5) | 2,480 (66.5) | 2,403 (67.3) |

### Statistical analysis

Data cleaning and analysis performed using STATA version 18. Exploration of the data was done to check for duplicates, unusual observations, and missing values prior to analysis. To deal with Missing values on the exposure variables multiple imputation methods were applied for the variables of less than 20% missing values. Some of the variables were re-categorized based on previous literature and plausibility (26).

The data was set to consider the complex nature of survey design by application of weights, primary sampling unit (cluster) and strata. Descriptive statistics were summarized using frequency and proportions for categorical. Since the prevalence of the adequate receiving of antenatal care services was greater than 10% so classical logistic regression was not fit for modelling and may overestimate or underestimate the results therefore, we used the alternative to logistic regression which fit model best based on the Akaike Information Criterion (AIC) and Bayesian Information Criterion (BIC). To determine factors associated with the receiving of the adequate ANC services the Poisson regression models were used as an alternative to logistic regression, the model estimated the prevalence risk (PR) and 95% confidence interval (CI) for improved precision of parameter estimates in accordance to other studies(31).Variables which were significant during crude analysis were considered in the adjusted analysis to determine the model which explain better the outcome of interest, then we created several adjusted analysis models to identify independent variables linked to receiving adequate ANC components.

## RESULTS

### Services received by socio-demographic characteristics

Table 1 highlights ANC services received by socio-demographic characteristics. Among 5,263 participants of this study, the mean (SD) age was 28.9(7.1). More than half (range 54.1%-56%) of study participants with primary education received ANC components. More than half (range 56.4%-57.7%) of the respondents who were married received ANC components. The majority of respondents (range 66.3%-67.8%) who received ANC services were not working. Around half of participants (range 46.7%-53.1%) had less than four ANC visits. Some of the study participants (range 32.4%-39.7%) who received ANC services were from poor households. Most of the study participants (range 65.5%-67.7%) whose partners education was primary received ANC components. Slightly less than half of the participants (range 44.2%-45.4%) who received ANC services were young adults (24 to 35 years).

### Proportion of adequacy of ANC services among WRA

In Tanzania, 41% of women of reproductive age (WRA) received sufficient ANC components. The rest, comprising the remaining percentage, did not attain this level of adequacy in their ANC reception. (Figure 1)

**Fig 1:**
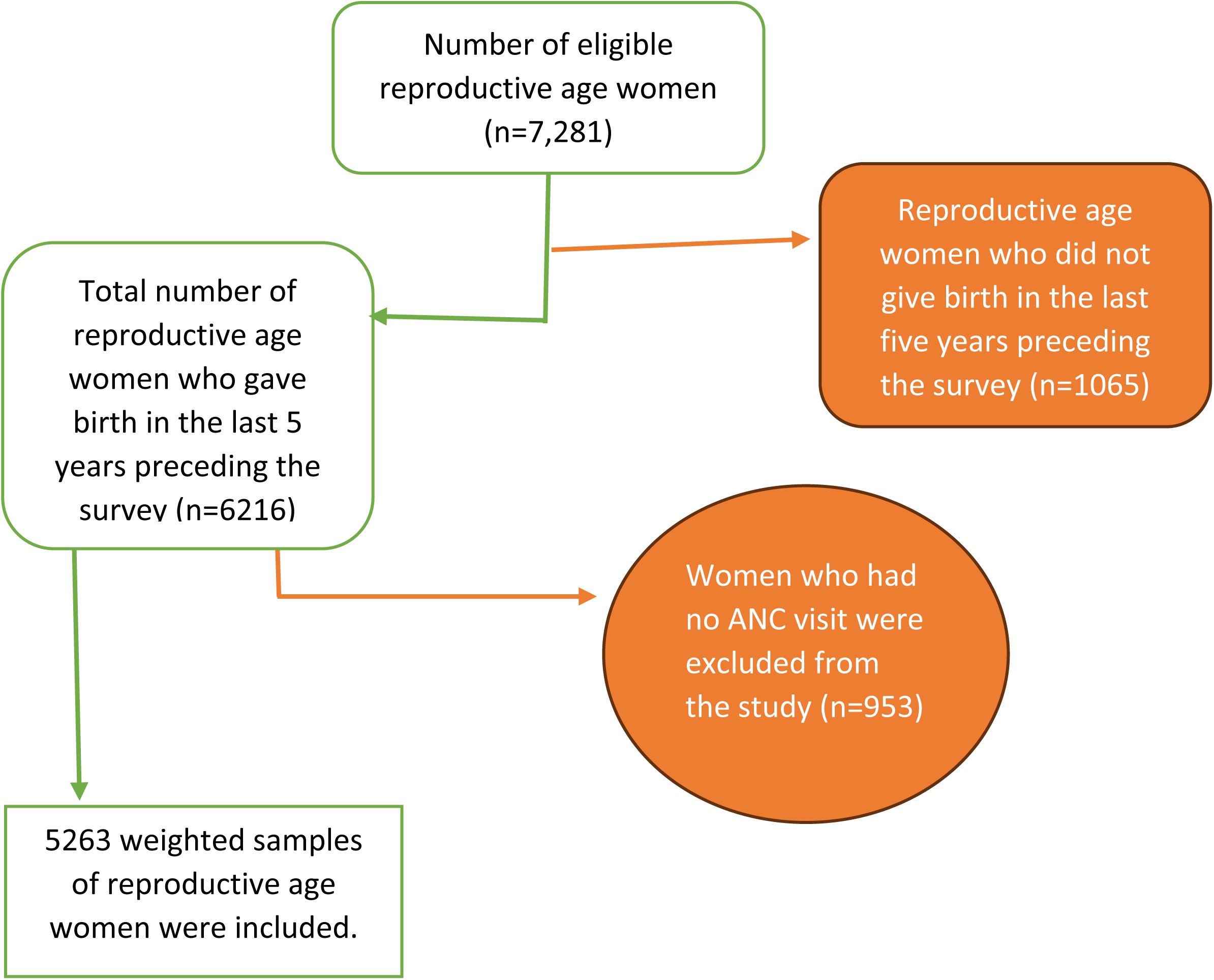
Schematic presentation showing the sampling and exclusion procedures to identify the final sample size in 2022 TDHS.

**Figure 1:**
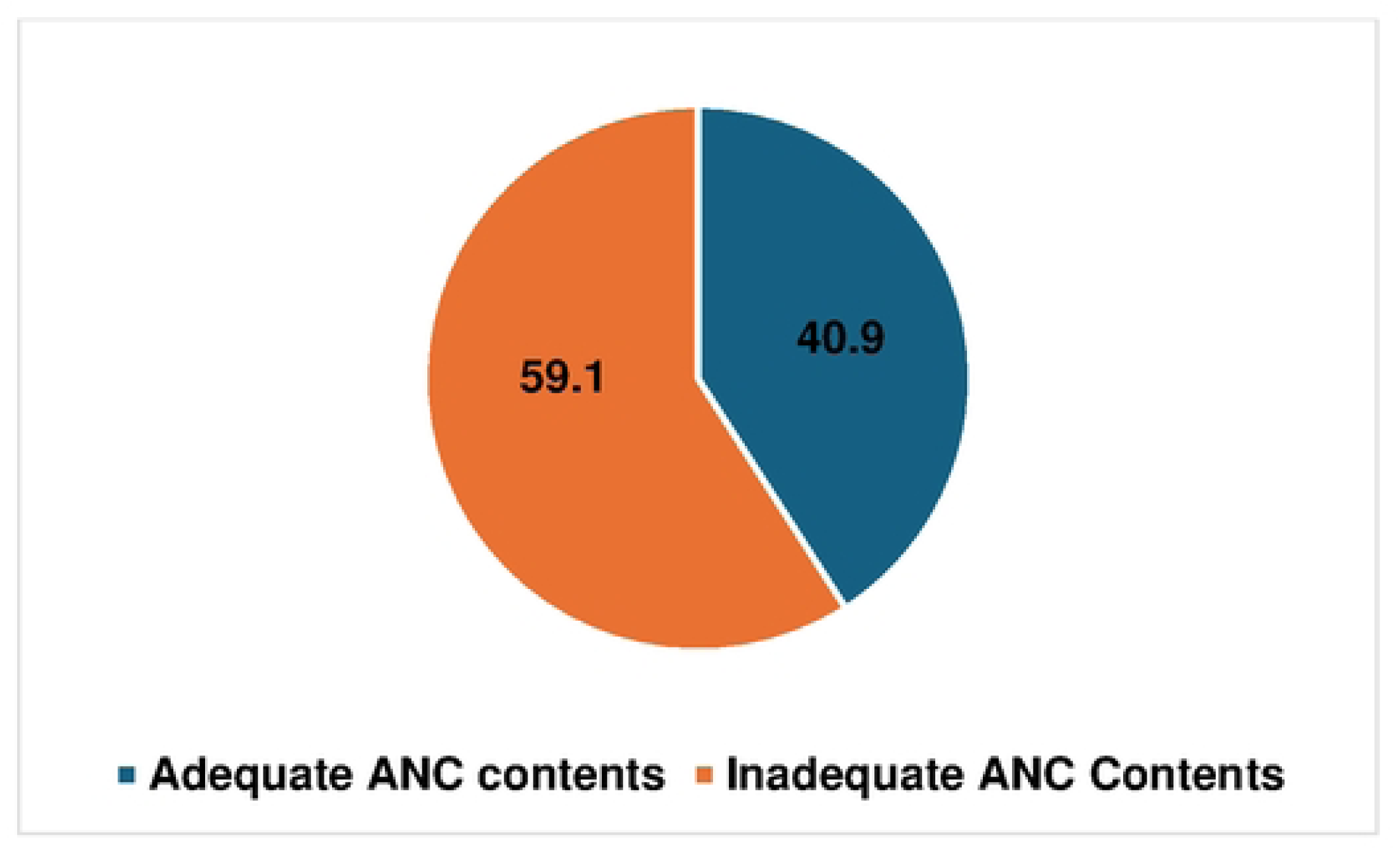
Proportion of adequacy and inadequacy reception of ANC services.

### ANC services coverage

Among women of reproductive age who received ANC for their most recent birth, the majority (77%) had their blood pressure measured, most of them (70.4%) had a urine sample taken, 91.6% had a blood sample taken, and almost all (96.1%) had their baby’s heartbeat checked. The majority (70.1%) of women received counselling on maternal diet and breastfeeding (70.1%), and approximately two-third (67.1%) were asked about vaginal bleeding (Figure 2).

**Figure 2:**
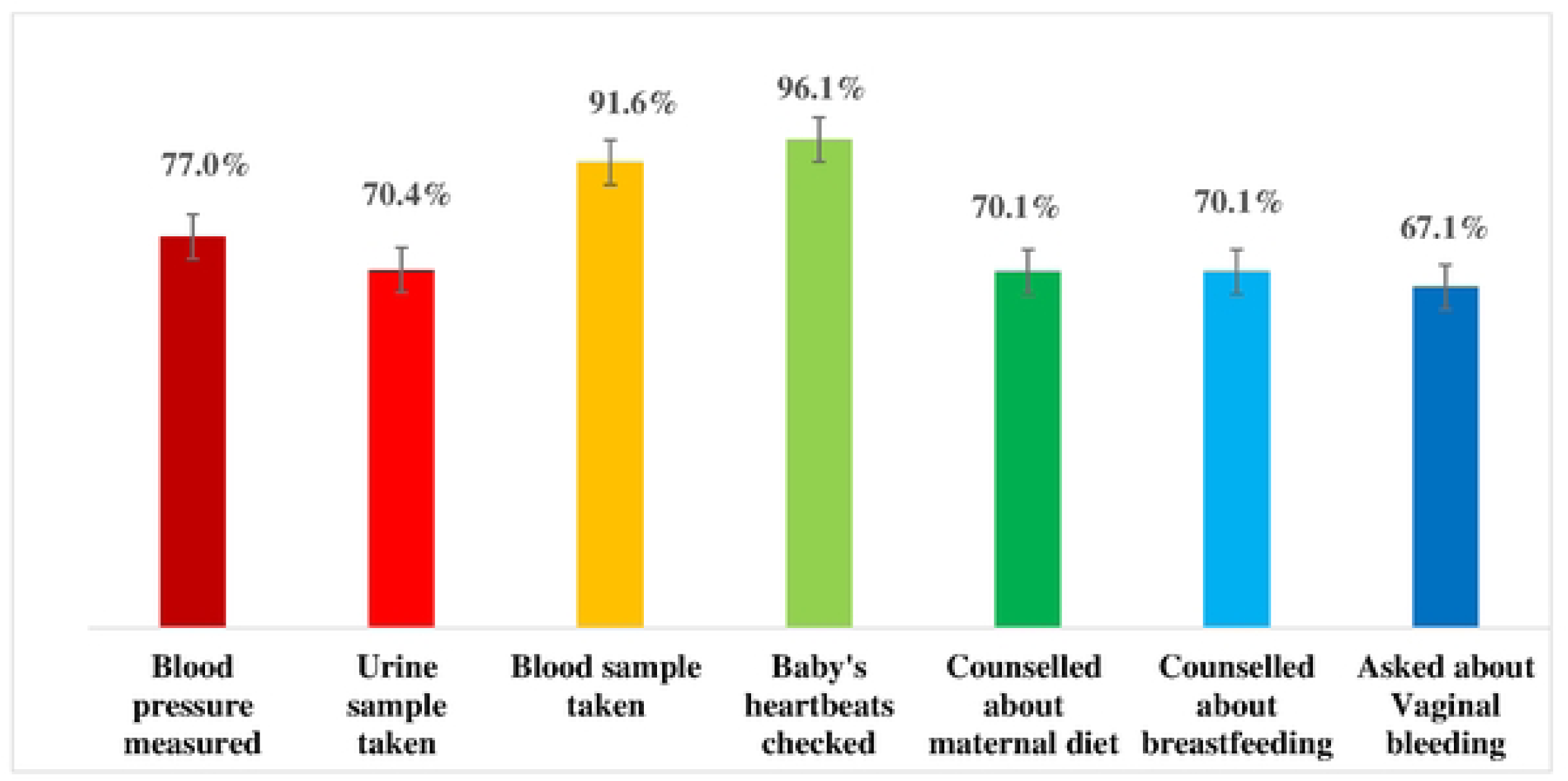
Services of antenatal care*(Among women who received ANC for their recent birth, the percentage with selected services)*

### Distribution patterns regarding the number of ANC services obtained by WRA in Tanzania

Figure 2 shows the number of ANC services received by WRA. More than two-fifth (40.9%) of study participants received all 7 ANC components. Almost one-fifth (16.3%) of the respondents received 6 ANC components. Few participants (14%) received 5 ANC components. More than one-tenth (12.1%) of the participants received 4 ANC components. Very few (8.4%) of the respondents received 3 ANC components. Among participants 5.8% and 2.3% of them received 2 and 1 ANC services respectively. Extremely few (0.5%) of the participants did not receive any ANC component. (Figure 3)

**Figure 3:**
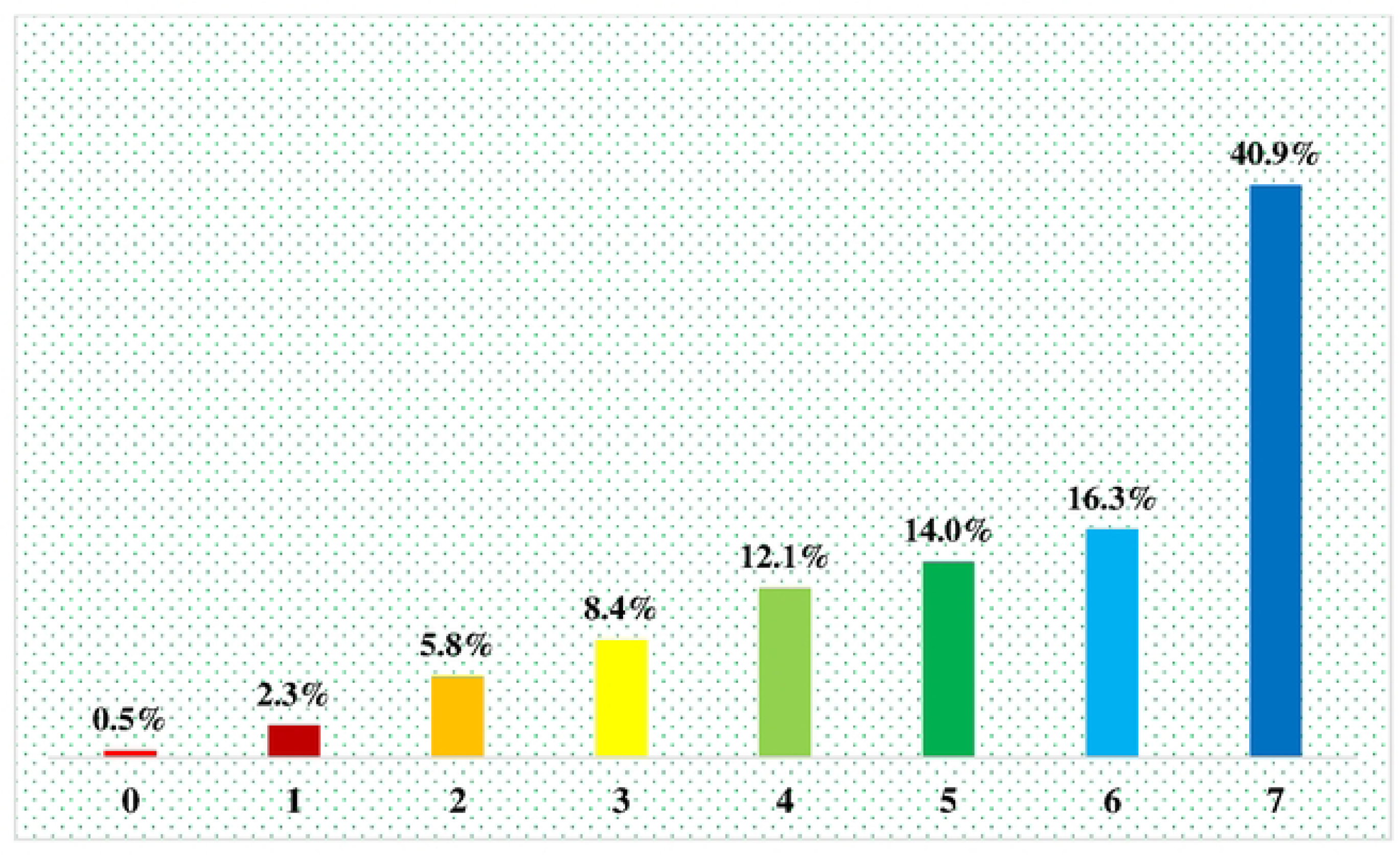
Distribution patterns regarding the number of ANC services obtained by WRA in Tanzania.

### Factors associated with adequate reception of ANC services

The factors that remained to be significant (p<0.05) associated with associated with adequate reception of ANC services in multivariable analysis among WRA in Tanzania were education, ANC visits, wealth index, birth order, age and residence. Women with secondary or higher education were more likely to receive adequate ANC services (RR= 1.42, 95% CI :1.22-1.66) compared to those with no education. Respondents who had more than 4 ANC visits were more likely to receive adequate ANC services (RR= 1.39, 95% CI :1.27-1.52) than those with less than 4 ANC visits. Higher prevalence of receiving adequate ANC was among women who were from rich house compared to those from poor household (RR= 1.42, 95% CI :1.22-1.66). Participants whose birth order was 2+, were 10% less likely (RR= 0.90, 95% CI: 0.82-0.99) to receive adequate ANC services than those whose birth order was 1. Older women aged 35 years and above were more likely to receive adequate ANC services (RR =1.35, 95% CI :1.21-1.50) compared to the younger participants aged 15-24. Women residing in rural areas were 18% less likely to receive adequate ANC services (RR =0.82, 95% CI :0.71-0.95) compared to those in urban (Table 2).

**Table 2:** Factors associated with adequate reception of ANC services among WRA in Tanzania 2022.

| Variable | Crude PR (95% CI) | Adjusted PR (95% CI) |
| --- | --- | --- |
| **Education** |  |  |
| No education | 1 | 1 |
| Primary | 1.51(1.28-1.78) | 1.27(1.09-1.50) |
| Secondary or higher | 2.19(1.28-2.58) | 1.42(1.22-1.66) |
| **Marital status** |  |  |
| Unmarried | 1 |  |
| Married | 0.97(0.89-1.06) |  |
| **Employment status** |  |  |
| Not working | 1 | 1 |
| Working | 0.89(0.80-0.98) | 0.91(0.82-1.00) |
| **ANC Visits** |  |  |
| <4 | 1 | 1 |
| >4 | 1.63(1.48-1.78) | 1.39(1.27-1.52) |
| **Wealth index** |  |  |
| Poor | 1 | 1 |
| Middle | 1.35(1.17-1.57) | 1.19(1.03-1.37) |
| Rich | 2.09(1.84-2.38) | 1.42(1.22-1.66) |
| **Partner's education** |  |  |
| No education | 1 | 1 |
| Primary | 1.47(1.15-1.87) | 1.09(0.85-1.38) |
| Secondary or higher | 2.18(1.71-2.79) | 1.19(0.94-1.51) |
| **Birth order** |  |  |
| 1 | 1 | 1 |
| 2+ | 0.77(0.69-0.86) | 0.90(0.82-0.99) |
| **Age categories** |  |  |
| 15-24 | 1 | 1 |
| 25-34 | 1.21(1.10-1.34) | 1.17(1.06-1.29) |
| 35-49 | 1.22(1.09-1.36) | 1.35(1.21-1.50) |
| **Residence** |  |  |
| Urban | 1 | 1 |
| Rural | 0.56(0.49-0.63) | 0.82(0.71-0.95) |

PR- Prevalence ratio

## Discussion

The study aimed to determine the proportion of ANC component coverage, factors associated with adequate reception of ANC services and distribution patterns regarding the number of ANC services obtained by WRA in Tanzania.

The study found that 41% of WRA had received adequate ANC component coverage with various factors such as education, wealth index, age, residence, number of ANC visits and birth order were significantly associated with uptake of ANC services among WRA in Tanzania.

Four out of ten women in Tanzania reported to have received adequate ANC component coverage, this indicate that more than a half of the study respondents have inadequate uptake of ANC services and this is in line with another researchers in Rwanda revealed that only one in ten women utilized all ANC components, indicating low and inadequate uptake of ANC components(32) whereas in Zambia, four out of ten women faced a similar issue(33). However among sub Saharan countries, a study done in Ghana revealed that seven out of ten women received adequate ANC components(34), this can be explained that early implementation of the free maternal health care policy since 2008 where women who enrolled on the National Health Insurance Scheme (NHIS) were having access to free maternal health services including ANC The study revealed that slightly less than half of the participants who received ANC services were young adults, this can be explained that most of the reproductive women enrolled in antenatal visits are at young adult age(35). This finding is in line with other studies done in Ethiopia(36) and Kenya(37) showing the critical significance of focusing on this young adults by employing an integrated approach not only to increase their utilization on ANC component but also awareness of various ANC services on this group as they are less experienced.

Furthermore, our study revealed rising trends in antenatal care (ANC) visits which corresponds to the increase in proportion of women receiving ANC components. We observed that the percentage of women receiving anywhere from zero to seven ANC services ranged from 0.5% to 41%, respectively. Interestingly, these findings differ slightly from a study conducted in Ethiopia, where the proportions of women receiving zero and eight services were 2% and 15%, respectively (9). This variation in findings could be to differences in sample size and study designs since our study covered the entire country, while the one done in Ethiopian had a localized focus.

Education had been found to have significantly associate with uptake of ANC components, in which women with secondary or higher education were more likely to receive adequate and adequate ANC services compared to those with primary or informal education. This could be because the educated women most likely have adequate knowledge on ANC services and understand the importance of early booking for ANC as well as attending the recommended four visits (38) hence they tend to value ANC services and will take advantage of this compared to women with lower education (39). This finding is in line with a study done in Ethiopia which revealed that higher education level of the woman being a significant factor in determining adequate ANC utilization(40). Apart from being a major drive of socio-economic status, education has a great chance to be empowered and improved health literacy(41) highlighting the vital role that educational initiatives play in raising health literacy, enabling women to prioritize and understand the need of prompt, all-inclusive ANC care, and ultimately improving the health of mothers and their children.

Age was another factor highlighted in the study, in which middle adult women and above were more likely to receive adequate ANC services compared to the young adult participants this findings is consistent with another study done in Ghana which revealed that odds of adequate ANC coverage were higher among older age groups compared to young adults (42), This can be explained, as the age of the mother increases, it might increases their knowledge, understandings, health literacy and experience of pregnancy and its related complications (43) once again emphasizing the role of experience and education on ANC component utilization as mothers age increases hence approaches that will include dynamic sharing and flow of experience among the two groups under guide of professional personnel could be explored.

Furthermore, respondents who had more than four antenatal visits had more likely to receive adequate ANC services than those with less than four ANC visits. In the same way as the previous studies done in in Myanmar(44) and Nigeria(27) . This can be due to adaptation of the current WHO guideline which recommends 8 number of ANC contacts/visit, as more frequent ANC visits increase opportunities to interact with health care workers and potentially enhance health talks and ANC in general hence enhancing adequate number of ANC visits should be in hand with human and material resources to ensure the best quality(27).

Respondents whose birth order was 2+, were less likely to receive adequate ANC services than those whose birth order was one or less than two, this study result consistent with another studies done in Kenya(37) and Zambia(45) and Haiti (46). This can be explained that high parity women have less desire to use recommended ANC visits that is also observed in Ethiopia due to the idea that they don’t require services since they had previous pregnancy and delivery experience may be the cause of this, less desire to use ANC leads to low ANC visits that results to poor ANC quality (47). This highlights the negative effect that experience can impose on ANC among high parity mothers calling the need to educate this group as experience doesn’t equate to ability handling of problems that can arise during pregnancy but only awareness hence avoiding the misinterpretation.

Women residing in rural areas were less likely to receive adequate ANC services compared to those residing in urban. This finding is consistent with another study conducted in sub-Saharan countries which found that women residing in urban areas were three times more likely to receive adequate ANC care than those in rural areas(33).But is inconsistent with study done in Canada which revealed no significant differences in uptake of ANC services among rural and urban residents(48) and Japan (49).

This could be due to difference in health sector policies, availability of adequate number of skilled health care personnels and infrastructural development between developed high-income countries like Canada and Japan compared to developing low middle income countries like Tanzania. Therefore, increasing the availability of trained healthcare workers and making numerous attempts to enhance accessibility particularly in primary care, can close the gap on the two groups’ adequate use of ANC components. Many rural communities lack readily accessible health care facilities, particularly those equipped to provide comprehensive antenatal care (ANC)(50).

Women who were from rich families/households were more likely to receive adequate ANC compared to those from poor household which is consistent with other study done in Zambia(51) . This can be clarified by stating that, participants in this study who lived on higher household incomes may have more resources available such as access to transportation or the ability to travel far distances to the ANC clinics within the areas with the expectation of receiving better quality care(52).

The strength of our study lies in its large sample size and nationwide coverage, ensuring representation across diverse demographics and geographic regions. This comprehensive approach enhances the generalizability of our findings to the broader population, making them valuable for government, policymakers, and stakeholders involved in maternal and child health initiatives. Moreover, our study stands as the first of its kind to thoroughly examine the factors influencing and distribution of adequate antenatal care services received in the country, addressing a significant information gap. However, it’s important to acknowledge the limitations inherent in our research design. Utilizing a cross-sectional approach means that data was collected at a single point in time, restricting our ability to establish causality between variables. Therefore, further qualitative research involving women, healthcare providers, and facilities is essential to gain deeper insights into the determinants of ANC component adequacy and the nuances of care delivery.

## Conclusion and recommendation

The overall findings indicate a significant gap in the uptake of antenatal care (ANC) services among women of reproductive age, with only 41% receiving adequate ANC coverage. Due to the findings from our study, we advocate for the strengthening of policies promoting eight or more ANC visits which could enhances the uptake of the recommended ANC components. Factors such as urban residence, four or more ANC visits, formal education, middle adult, and higher wealth status significantly influenced ANC component uptake. Addressing these disparities requires expanding health insurance coverage and poverty reduction policies. Integrated approaches involving health care facilities and communities, as well as innovative strategies targeting young adults, are recommended to improve ANC access and utilization. Strengthening the policy of four or more ANC visit with regular monitoring and data collection are also crucial for aligning ANC services with current WHO guidelines and improving overall ANC contact and utilization rates.

## Data Availability

The data underlying the results presented in the study are available from http://www.dhsprogram.com

## Acknowledgments

The authors are grateful to MEASURE DHS for providing them with the data set.

## Funding

This research did not receive any specific grant from funding agencies in the public, commercial, or not-for-profit sectors.

## Disclosure

The authors report no conflicts of interest in this work.

## Abbreviations

AIC: Akaike Information Criteria
ANC: Antenatal Care
BIC: Bayesian Information Criteria
CI: Confidence Interval
DHS: Demographic and Health Survey
EA: Enumeration Area
MMR: Maternal Mortality Ration
GDM: Gestational Diabetes Mellitus
NBS: National Bureau of Statistics
PR: Prevalence Risk
PSU: Primary Sampling Unit
WRA: Women of Reproductive Age
TDHS: Tanzania Demographic and Health Survey

## Authorship contributions

Jovin R Tibenderana and Sanun Ally Kessy: Conceptualization, Data curation, Formal analysis, Methodology, Visualization, Writing – original draft, Writing – review & editing. Ndinagwe Lloyd Mwaitete and John Elyas Mteng: Writing – original draft, Writing – review & editing, Visualization. Jomo Gimonge and Dosanto Felix Mlaponi: Writing – review & editing, Visualization. Fabiola Moshi: Writing – review & editing, Supervision.

## Notes

### Competing Interest Statement

The authors have declared no competing interest.

### Funding Statement

The author(s) received no specific funding for this work.

### Author Declarations

The formal written request was submitted to the DHS program and approval was given to access and utilize data from http://www.dhsprogram.com. The questionnaire for standard DHS was reviewed and approved Medical Research Council of Tanzania and the Zanzibar Health Research Institute and ICF’s Internal Review Board (IRB). Participants provided either written or verbal informed consent before participating in the survey.

